# Atypical intrinsic neural timescale in the left angular gyrus in Alzheimer’s disease

**DOI:** 10.1101/2023.06.12.23291278

**Authors:** Shota A. Murai, Tatsuo Mano, Jerome N. Sanes, Takamitsu Watanabe

**Author notes:** **Corresponding authors** S.A.M., International Research Centre for Neurointelligence (WPI-IRCN), The University of Tokyo Institutes for Advanced Study., Address: 7-3-1 Hongo Bunkyo-ku, Tokyo 113-0033 Japan., T.W., International Research Centre for Neurointelligence (WPI-IRCN), The University of Tokyo Institutes for Advanced Study., Address: 7-3-1 Hongo Bunkyo-ku, Tokyo 113-0033 Japan. **Competing interests** The authors declare that no competing interests exist.

## Abstract

Alzheimer’s disease (AD) is characterised by cognitive impairment and progressive brain atrophy. Recent human neuroimaging studies showed that such AD symptoms are linked with anatomical and functional changes seen in the default mode network (DMN), but the key brain region whose atrophy disturbs the neural activity of the DMN and consequently contributes to the symptoms of AD remains unclear. Here, we examined the intrinsic neural timescales (INT) of the regions in the DMN aiming to identify such a crucial brain area. We investigated INT since prior work demonstrated its utility in quantifying local neural dynamics and bridging knowledge gaps between atypical local neuroanatomical features and symptoms in neuropsychiatric disorders. We used resting-state functional MRI data and compared INT between individuals diagnosed with AD and age-/sex-/handedness-matched cognitively normal individuals. To exclude the possibility that a region outside of the DMN has such a critical role, we first performed an exploratory whole-brain analysis and found that only the left angular gyrus, a region within the DMN, exhibited a shorter INT in the AD group compared to those with normal cognition. We also identified an AD-specific decrease in the grey matter volume in the left angular gyrus and revealed that such regional atrophy shortened the INT of the angular gyrus and consequently reduced the overall INT of the DMN. Moreover, we found that the overall shorter DMN INT led to the symptoms of AD, particularly, its impairments of attention control. Taken together, our findings indicate that the left angular gyrus serves as a key brain region whose structural atrophy and resultantly shorter INT destabilise DMN neural dynamics and contribute substantially to AD cognitive decline. Clinically, the outcome of this study indicates that INT of the left angular gyrus could serve as a convenient biomarker for the cognitive symptoms of AD.

## Introduction

Alzheimer’s disease (AD) is the most common form of dementia, ostensibly due to progressive brain atrophy. In both human and non-human animal model systems, pathological findings show that cognitive impairment in AD and its neural degeneration are specifically associated with amyloid and tau depositions in myriad brain regions, including the medial and lateral temporal lobe and several parietal regions^1-7^.

Neural degenerative changes in AD commonly occur in regions within the so-called default mode network (DMN)^8,9^, including the medial prefrontal cortex, precuneus and angular gyrus^4,10-15^, and the atrophy seen in some of these DMN regions is specific to AD^12-14^.

Moreover, AD yields atypical activity in the DMN as assessed by resting-state functional magnetic resonance imaging (rsfMRI)^16,17^: specifically, AD patients showed significantly weaker functional connectivity within the entire DMN^16,18^ or only its posterior part^19,20^, and such atypical functional connectivity in the DMN was associated with amyloid^4,19,21^ and tau deposition^22-24^ within the DMN’s brain regions. Given the DMN’s crucial role in cognitive capability in older adults^25^, these findings led us to hypothesise that the anatomical changes within the DMN would disturb its neural dynamics, which then might lead to the cognitive impairment of AD.

However, it remains unclear whether one or more brain regions in the DMN have such a critical role in the pathophysiology of AD. Anatomically, multiple neuroimaging studies described AD-specific atrophy in the temporal-parietal junction^12-14^, but whether such focal atrophy in the DMN affects the network’s neural activity in AD was not examined. Several functional MRI studies with AD patients identified atypical neural dynamics in parietal regions within the DMN^19,20,26^; but these studies did not directly test whether such aberrant neural activation was attributable to local brain atrophy.

Consequently, the DMN region whose focal anatomical change induces network-level neural dysfunction that leads to AD symptoms in humans has not yet been identified.

Here, we examined the “intrinsic neural timescale” (INT) of resting-state neural activity for all brain regions^27-29^ aiming to identify such trigger neural zones that are crucial for the cognitive decline in AD.

INT, also known as temporal receptive windows^30-35^ or temporal receptive fields^36^, represents the time period during which a brain region integrates neural inputs from other regions^37^. Regions within the DMN and frontal-parietal network typically exhibit longer INT compared to other brain regions, which enables the DMN to incorporate diverse neural inputs from various remote brain areas into their intrinsic signal processing^30-33,36,38-40^. In contrast, sensory-related cortices, such as the primary visual cortex, typically exhibit a shorter INT, which permits these regions to respond to their inputs with higher temporal fidelity^30,38,41,42^.

In addition to such functional findings, we previously showed that this temporal index for local neural activity is related to neuroanatomical features: a brain region with a larger grey matter volume (GMV) more likely had a longer INT^27^. This relationship between INT and GMV has biological relevance insofar as a brain area with a higher neuronal density, as proxied by a larger GMV, tends to have more recursive neural processing, which prolongs the autocorrelation of its intrinsic neural activities, thereby yielding a longer INT.

Taken together, these previous findings indicate the possibility that INT could link local neuroanatomical changes to atypical neural activity and symptomatic behaviours in neurological and neuropsychiatric disorders^27^.

Given this background, we focused on INT aiming to identify a key DMN region whose atrophy affected its INT, which in turn then disturbed DMN neural dynamics, which then potentially led to AD symptoms. By analysing rsfMRI data obtained from AD patients and matched healthy controls, we tested the following four working hypotheses: (i) one or more DMN regions should exhibit a significantly shorter INT in people with AD; (ii) the overall INT of the entire DMN should also be shortened in AD individuals, and the DMN regions with shorter INT should have atypically smaller GMV; (iii) the smaller GMV of the specific DMN regions should induce the shorter INT of the regions, which results in shortened overall INT of the DMN; (iv) the shorter INT of the DMN regions or overall DMN should be associated with AD symptoms.

## Materials and methods

### Participants

We used an MRI dataset collected by the Washington University Knight Alzheimer Disease Research Center^43^, which has public availability as the Open Access Series of Imaging Studies (OASIS-3)^44^. The data acquisition procedures from people diagnosed with AD and matched healthy controls received ethics approval for human subject research by the Washington University Human Research Protection Office, and all participants or their guardians provided written consent.

Participants completed clinical assessment protocols in accordance with the Uniform Data Set (UDS)^45^, which included medical history, physical examination, neuropsychological test battery and neurological evaluation. The diagnosis of AD was made based on the Clinical Dementia Rating scale (CDR)^46,47^: individuals with more than a zero CDR score were diagnosed as having AD (‘AD dementia’), while those with zero CDR score were labelled as non-demented (‘cognitively normal’, CN)^43,44,48^.

Aiming to reduce within-group heterogeneity, we excluded AD individuals who showed relatively normal scores in the Mini-Mental State Examination (MMSE), instead focusing on AD individuals with MMSE scores ≤23^49,50^.

We also excluded AD participants whose clinical assessment occurred ≥365 days before the MRI scan, whose head motion during scanning exceeded 3 mm, or who had dementia-relevant cardiovascular diseases or strokes. The control group included age-/sex-/handedness-matched adults. With these criteria, we included 32 individuals with AD and 138 CN individuals (Table 1).

**Table 1.**
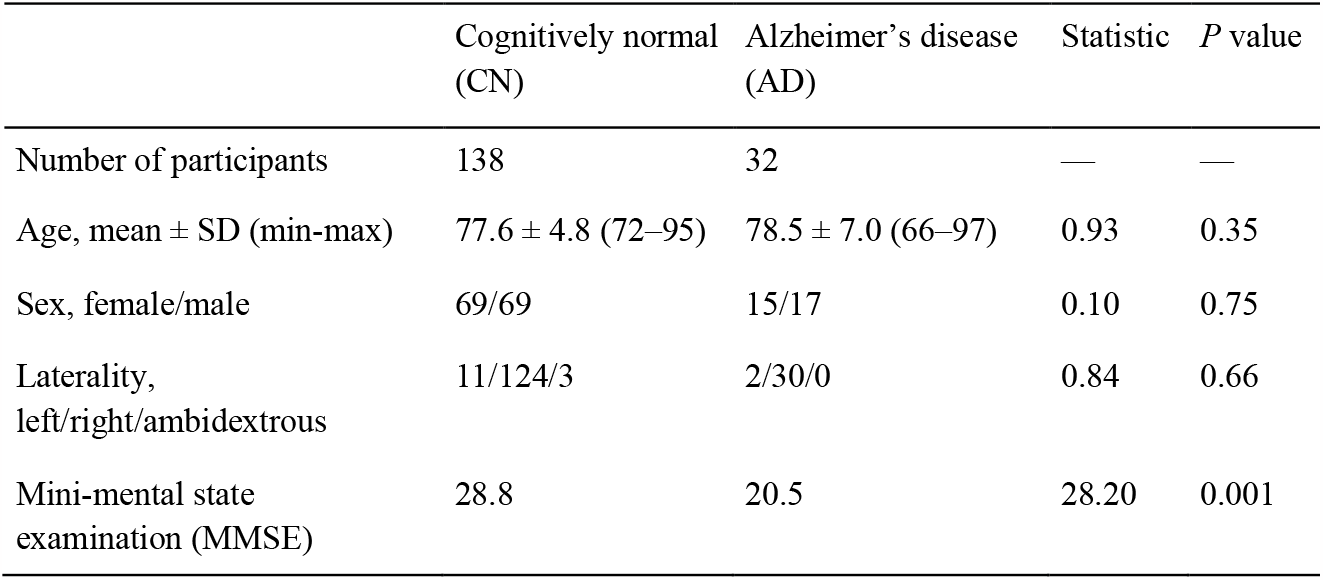
Demographic data.

### MRI data acquisition

The rsfMRI and anatomical MRI data were collected with a 3T MRI scanner (TIM Trio, Siemens) using a 20-channel head coil. The rsfMRI data were acquired using a multi-band echo-planar imaging sequence in a single run lasting ∼6 min (repetition time = 2.2 s, echo time = 27 ms, 36 slices, interleaved, flip angle 90°, in plane resolution = 4 × 4 mm^2^, slice thickness = 4 mm); for anatomical MRI, structural images were obtained using T1-weighted sequence (repetition time = 2.4 s, echo time = 3.16 ms, flip angle = 8°, in plane resolution = 1 × 1 mm^2^, slice thickness = 1 mm). The participants were asked to lie quietly with their eyes open during the rsfMRI scanning.

### Preprocessing of rsfMRI data

SPM12 (www.fil.ucl.ac.uk/spm) after discarding the first five volumes. This preprocessing procedure consisted of realignment, unwarping, slice-timing correction and normalisation to a standard template (ICBM 152). We then performed regression analyses to remove the effects of head motion, white matter signals and cerebrospinal fluid signals, and finally conducted band-pass temporal filtering (0.01–0.1 Hz). Note that participants who had high head motion (mean motion >3 mm) were excluded, which resulted in no significant difference in the mean head motion (*P* > 0.1 in a two-sample *t*-test) or maximum/mean framewise displacement (*P* > 0.2 in a two-sample *t*-test) between the AD and CN groups.

### Explanatory whole-brain analysis of INT

Using the preprocessed rsfMRI data, we evaluated INT for all brain regions at a single-participant level following previously used procedures^27^. First, an autocorrelation function (ACF) of the preprocessed rsfMRI signal (time bin = TR) was estimated, and the area under the curve of the ACF values was calculated within the initial period during which the ACF showed positive values. The point at which the ACF first reached zero was the upper limit of the initial period. After repeating this procedure for every voxel, we smoothed the resulting whole-brain INT map (Gaussian kernel, full-width at half maximum = 8 mm).

Using these INT maps, we estimated the group-mean maps by averaging them within groups and then compared the AD map to the CN map, using a random-effects model with age and sex information included as nuisance variables, setting a statistical threshold at *P*_FWE_ = 0.05 at the cluster level.

### INT and AD symptoms

For the brain areas showing significantly different INT between the AD and CN groups, we examined associations between their INT and the severity of AD symptoms. We calculated Pearson correlation coefficients between the average INT in these regions of interest (ROIs) and the MMSE scores in the AD group. The ROIs were defined as a 4mm-radius sphere centred at the peak coordinates found in the above whole-brain analysis, and an INT for each ROI was given as the average within the ROI. The effects of multiple comparisons between these ROIs were corrected using the Bonferroni correction. For these data and other statistical tests, we report exact probabilities from *P* = 0.05 to *P* =.0001 and use the less-than-equal symbol for *P* values less than 0.001.

### INT of DMN

We investigated INT within the entire DMN since we hypothesised a critical role of the DMN in AD and also since the DMN regions emerged in the above-noted exploratory analysis (see Results). We defined the DMN based on a functionally-determined brain parcellation system^51^.

We first calculated INT within the DMN by averaging INT, within-subjects, for all rsfMRI time-series data across all the regions within the DMN. We next assessed whether INT of the ROIs emerging from the exploratory analysis (see Results) correlated with the overall DMN INT.

### Regional GMV and INT

We also calculated the GMV of the ROIs that were detected in the above INT analysis. We estimated the GMV based on T1-weighted MRI images using SPM12 for each participant as follows. First, the MRI images were segmented into grey matter, white matter, and cerebrospinal fluid with the New Segment Toolbox^52^. Second, using the DARTEL Toolbox^53^, the segmented grey matter images were aligned, warped to a template space, resampled to 1.5-mm isotropic voxels and registered to a participant-specific template. The grey matter images were then smoothed with a Gaussian kernel (full-width at half maximum = 8 mm). Using these preprocessed T1-weighted images, we then calculated the GMV of the ROI that was detected in the above analysis. Next, we examined a Pearson correlation coefficient between the GMV and INT in the ROIs identified in the INT analysis.

### Mediation effect of regional INT

To assess potential causal relationships between the local GMV, local INT and DMN INT, we then conducted a nonparametric mediation analysis. In the analysis, we set the independent variable using the GMV of the ROIs found in the exploratory whole-brain analysis and incorporated DMN INT as a dependent variable. The INT of the ROIs were used as a mediator variable. The statistical significance of the mediation effect was calculated using a bootstrapping method.

As a control, we also performed the same mediation analysis using the GMV and INT of the brain regions that were found in the above-mentioned exploratory whole-brain analysis but showed only a marginally significant correlation between its INT and AD symptoms.

As another control, we also conducted the same mediation analysis using the GMV and INT of the brain regions in the other hemisphere compared to the original ROIs that were identified in the whole-brain explanatory analysis and correlation test with AD symptoms.

### Associations with AD symptoms

Next, we examined the causal relationships between the local INT, DMN INT and AD symptoms using a nonparametric mediation analysis. Before this analysis, we confirmed whether the DMN INT correlated with the AD symptoms.

We conducted two types of mediation analysis to test the following two models: (i) the atypically shorter INT of the ROIs found in the exploratory whole-brain analysis shortens DMN INT, which results in the AD symptoms; and (ii) the atypicality of the ROI INT is strong enough to account for the link between DMN INT and AD symptoms.

To test model (i), we set the ROI INT as an independent variable and defined the mediator variable using DMN INT. To test model (ii), we used DMN INT as an independent variable and adopted the ROI INT as a mediator variable. In both cases, the MMSE scores indicating the AD symptoms were set as a dependent variable.

After determining which model worked in the current dataset, we examined the specificity of the DMN in this brain-behaviour relationship. For this positive control test, we conducted another mediation analysis after replacing DMN INT with INT of the other six brain networks defined in the same brain parcellation system^51^ (i.e., frontal-parietal network, dorsal attention network, ventral attention network, limbic system, sensory-motor network and visual network).

### Associations with different cognitive components

We also searched for cognitive components that were specifically affected by the changes in DMN INT. To this end, we conducted a regression analysis between DMN INT and the scores of the following neuropsychological tests^45,54^: ‘Digit Span Test ‘for the measurement of attention; ‘Trail Making Test Part A ‘and ‘Digit Symbol Test ‘to evaluate processing speed; ‘Trail Making Test Part B ‘to quantify executive function; ‘Logical Memory Test ‘to measure episodic memory;

‘Boston Naming Test ‘and ‘Category Fluency Test ‘to evaluate language function. All the scores were converted to z-scores. The scores of Trail Making Tests were inverted to represent that a higher value indicated better cognitive performance.

## Results

### Whole-brain analysis of INT

We first performed an exploratory whole-brain analysis of INT (Fig. 1A) to identify brain regions that showed an atypical INT in the AD individuals compared to the age-/sex-/handedness-matched controls (CN; Fig. 1B and Table 1). We found that the AD group yielded significantly shorter INT compared to CN in the left angular gyrus (AG) and right supramarginal gyrus (SMG) (*P* < 0.001 for the left AG, *P* = 0.002 for the right SMG, *P*_FWE_ < 0.05; Fig. 1C and Table 2). No region showed a significantly longer INT in the AD individuals compared to the controls.

**Table 2.** Results of whole-brain analysis of intrinsic neural timescale.

|  |  |  | MNI coordinates |  |  | Cluster size |  |
| --- | --- | --- | --- | --- | --- | --- | --- |
|  | Region | Laterality | x | y | z | (voxels) | t-value |
| CN > AD | Angular gyrus | Left | -34 | -70 | 34 | 3466 | 5.15 |
|  | Supramarginal gyrus | Right | 60 | -38 | 30 | 1383 | 4.38 |

**Figure 1.**
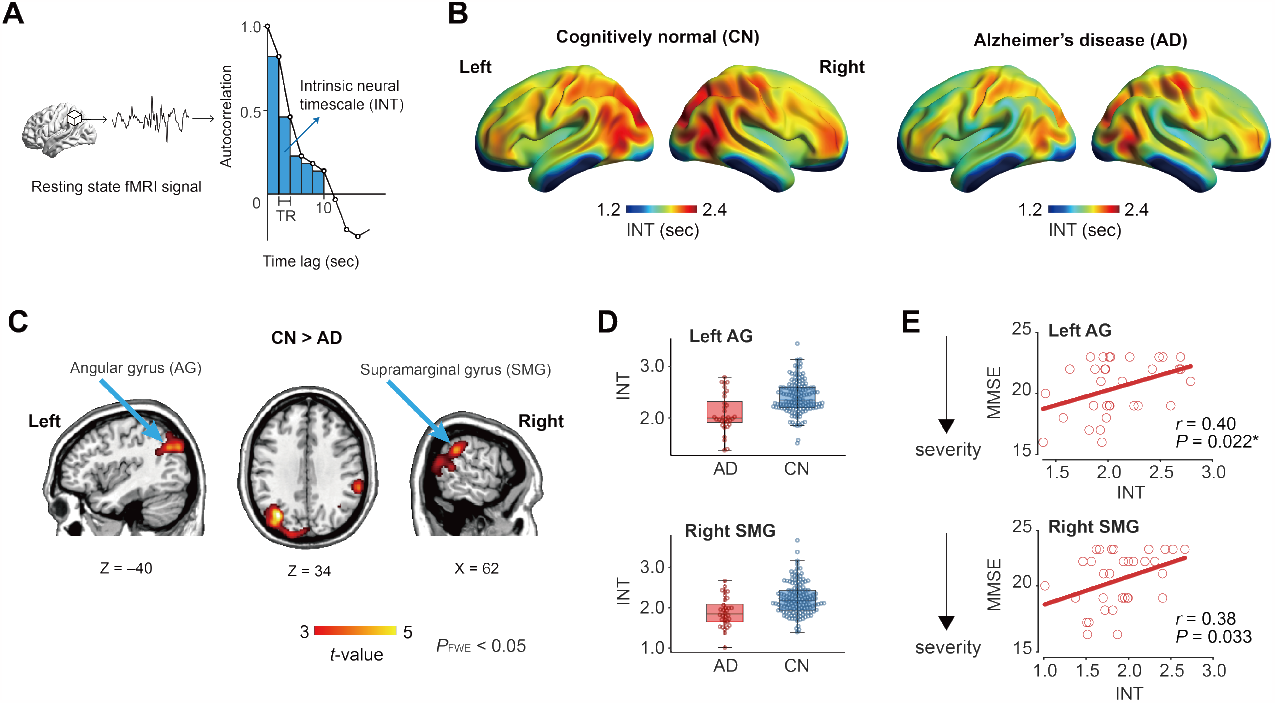
Intrinsic neural timescale and cognitive impairment. **(A)** To estimate an intrinsic neural timescale (INT), we calculated the area under curve of the autocorrelation function of each local rsfMRI signal. **(B)** Exploratory whole-brain analyses showed longer INT in the frontoparietal areas in both the cognitively normal (CN) and Alzheimer’s disease (AD) groups. **(C & D)** A voxel-wise comparison of INT between the CN and AD groups showed that the AD individuals had significantly shorter INT in the left angular gyrus (AG) and the right supramarginal gyrus (SMG, *P*_FWE_ < 0.05). No region showed a significantly longer INT in the AD group. Panel D shows INT of those regions for visualisation purposes. **(E)** In the AD group, a significant positive correlation between INT and MMSE was found in the left AG but not in the right SMG. Each circle represents each AD individual. *, *P*_Bonferroni_ < 0.05.

We then assessed potential associations between the severity of AD and the atypical INT observed in the left AG and right SMG. We found positive correlations between the MMSE and INT in both the left AG (*r* = 0.40, *P* = 0.022; Fig. 1E, upper panel) and the right SMG (*r* = 0.38, *P* = 0.033; Fig. 1E, lower panel); however, only the correlation seen in the left AG survived the Bonferroni correction for multiple comparisons. Given these results, we focused only on the left AG in subsequent analyses.

### INT of DMN

Next, we examined whether the shorter INT with the AG filled the gap between anatomical changes of the AG and the atypical activity of the DMN. We investigated DMN activity because, as stated in the Introduction, we hypothesised that atypical neural dynamics of the DMN should be closely related to AD symptoms and also because the left AG is a component of the DMN (Fig. 2A).

**Figure 2.**
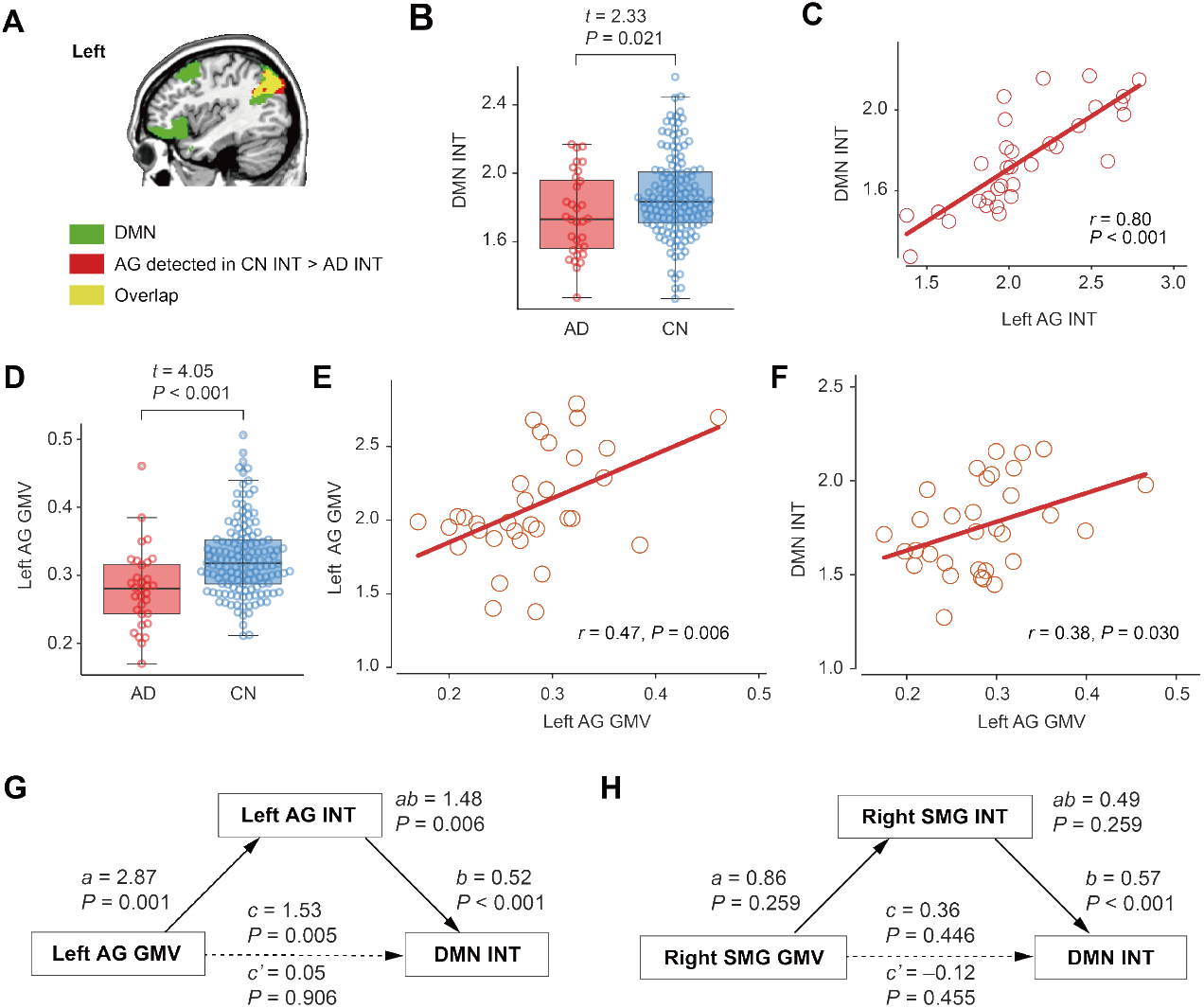
Intrinsic neural timescale of default mode network. **(A)** The default mode network (DMN, green) partially overlapped (yellow) with the left AG that showed a shorter INT in the AD group relative to the CN group (red). **(B)** INT of the DMN was significantly shorter in the AD group compared to the control. **(C)** AG INT was correlated with DMN INT in the AD group. Each circle represents an AD individual. **(D)** The GMV of the left AG was smaller in the AD group than in the CN group. **(E)** In the AD individuals, AG GMV was significantly correlated with AG INT. **(F)** In the AD group, AG GMV was also correlated with DMN INT. Each circle represents an individual. **(G)** A nonparametric mediation analysis showed that the atypically reduced AG GMV shortened AG INT, which resulted in shorter DMN INT. *a* denotes a regression coefficient of AG GMV on AG INT, and *b* indicates that of AG INT on DMN INT. **(H)** We did not find such a significant mediation effect for the right SMG, which was found in the exploratory whole-brain analysis of INT (Fig. 1C) and showed a marginal association with AD symptoms (Fig. 1E). *c* represents the direct effect of AG GMV on DMN INT without the mediator effect, whereas *c’* represents that with the mediator effect of AG INT. *ab* indicates the indirect effect of AG GMV on DMN INT via AG INT.

To address this hypothesis, we first calculated the INT of the DMN and confirmed that DMN INT was significantly shorter in AD compared to matched controls (*t*_168_ = 2.33, *P* = 0.021, η^2^= 0.03 in a two-sample *t*-test; Fig. 2B). In addition, we found that DMN INT correlated with AG INT in the AD group (*r* = 0.80, *P* ≤ 0.001; Fig. 2C).

### GMV of AG

To continue testing our hypotheses, we then calculated the GMV of the left AG and confirmed that AG GMV was significantly reduced in the AD group compared to that in the CN group (*t*_168_ = 4.05, *P* ≤ 0.001, η^2^= 0.09 in a two-sample *t*-test; Fig. 2D). Additionally, left AG GMV correlated with left AG INT in the AD group (*r* = 0.47, *P* = 0.006; Fig. 2E). This GMV-INT association is consistent with our previous finding^27^.

### Mediation effect of AG INT

Using these aforementioned estimations, we then performed a nonparametric mediation analysis and examined the extent to which left AG INT explained the seemingly significant direct correlation between left AG GMV and DMN INT (*r* = 0.38, *P* = 0.03; Fig. 2F).

We found that the total effect of left AG GMV on DMN INT (*c* = 1.53, *P* = 0.005; Fig. 2G) was entirely explained by a mediation effect of left AG INT (*c’* = 0.05, *P* = 0.09; *ab* = 1.48, *P* = 0.006). That is, this complete mediation model suggests that the smaller GMV of the left AG reduces left AG INT, which results in the shorter DMN INT.

To assess the specificity of the left AG GMV on DMN neural dynamics, we assessed whether GMV and/or INT of the other DMN regions mediated the total DMN INT. Since INT in the right SMG differed between the AD and CN groups (Fig. 1D) and showed a marginally significant correlation with cognitive decline in the AD individuals (Fig. 1E), we reasoned that SMG INT might also exert a mediation effect on the relationship between SMG GMV and DMN INT. However, this analysis failed to show such a significant mediation effect (*P* = 0.26; Fig. 2H). Moreover, we confirmed that INT of the right AG, another DMN region and counterpart of the left AG, did not have a significant mediation effect on the relationship between GMV of the right AG and DMN INT (*P* = 0.75). We also found no significant mediation effect for any of the other DMN regions (superior medial prefrontal gyrus, anterior cingulate, bilateral superior frontal gyri, bilateral inferior temporal gyri, bilateral parahippocampal gyri and middle cingulate). These negative findings suggest that the left AG has a specific role in linking its local anatomical change to the overall DMN activity in AD individuals.

### Association with AD symptoms

We next investigated whether the atypically shorter INT occurring in both the left AG and overall DMN in AD patients could explain AD symptoms. Since both left AG INT and DMN INT correlated with AD symptoms (Fig. 1E for left AG INT and Fig. 3A for DMN INT), we tested the following two models to understand the interrelationships among left AG INT, DMN INT and AD symptoms, as indexed by the MMSE test. First, we assessed whether atypically shorter left AG INT reduces DMN INT and consequently contributes to AD symptoms, and second, whether the shortened left AG INT is so substantial that it can explain the association between DMN INT and AD symptoms.

**Figure 3.**
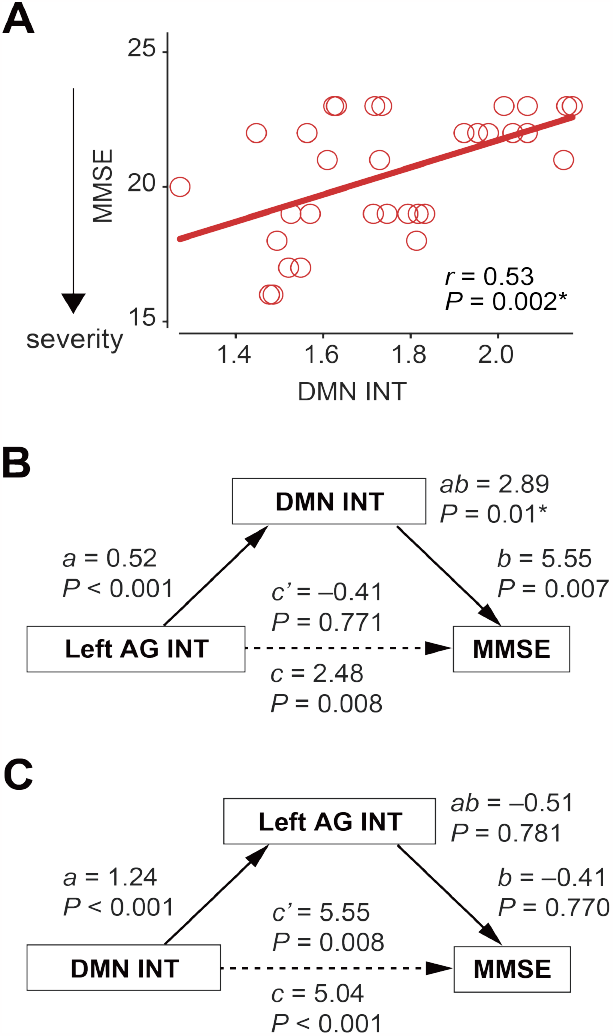
Association between intrinsic neural timescale and grey matter volume in left angular gyrus. **(A)** In the AD individuals, INT of the DMN was correlated with their AD-related symptoms. **(B)** In the AD group, a non-parametric mediation analysis showed that DMN INT accounts for the influence of AG INT on cognitive impairments of AD. * indicates *P*_Bonferroni_ < 0.05 between the two models. **(C)** A separate mediation analysis suggests that AG INT cannot explain the association between DMN INT and AD symptoms.

We examined these two models using a mediation analysis and only found support for the first model. That is, DMN INT mediated the entire relationship between left AG INT and AD symptoms (*P* = 0.01; Fig. 3B). By contrast, left AG INT did not account for the significant direct association between reduced DMN INT of AD patients and their symptoms (*P* = 0.78; Fig. 3C).

To control for the possibility that DMN INT simply reflected a generalised decrease in neural dynamics across all brain regions, we assessed whether the INT of brain networks other than the DMN mediated the direct effect of left AG INT on the MMSE score. We found that, except for the DMN, none of the brain networks exhibited atypical INT that could explain the direct influence of the left AG INT on the MMSE score (mediation effects, *P* > 0.08). These null results provide further evidence that the DMN has a specific functional role in bridging the gap between atypical neural processing in the left AG and AD symptoms.

Considering this result (Fig. 3B) along with other findings (Fig. 2G), we infer that, in the AD patients, their atypical reduction in GMV of the left AG shortened its INT, shorter AG INT reduced DMN INT, and finally that the shorter DMN INT led to their cognitive impairment.

### Specific cognitive components

Finally, we assessed whether DMN INT exerted specific effects on cognitive components in AD (Fig. 4). We found that DMN INT correlated significantly in the AD patients only with attention (*R*^2^ = 0.28, *P* = 0.002) and not with processing speed, executive function, memory, or language (*P* > 0.01). These results suggest that the alteration of DMN INT in AD has a specific effect on attention-related symptoms.

**Figure 4.**
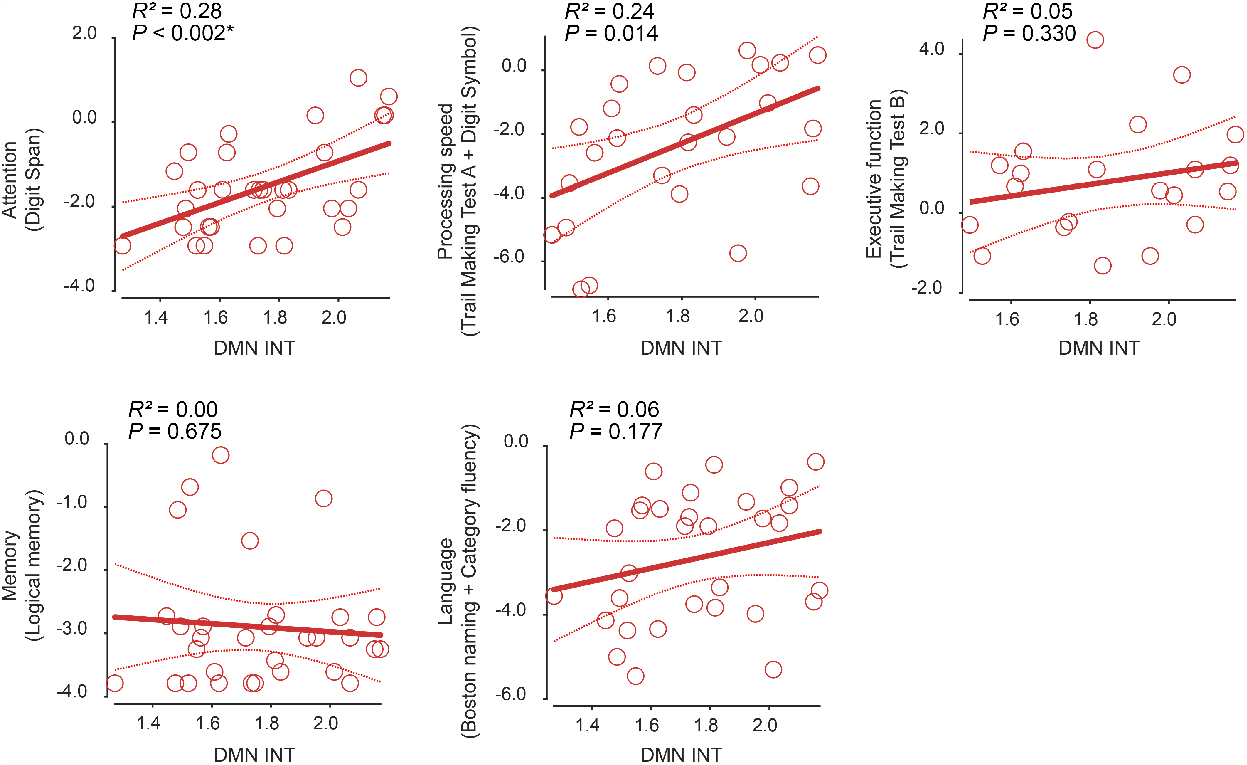
Associations between intrinsic neural timescale of default mode network and different cognitive components. In the AD group, DMN INT was significantly associated with attention but neither with processing speed, executive function, memory nor language. Each circle represents an individual. * indicates *P*_Bonferroni_ < 0.05 between the five cognitive components.

## Discussion

By focusing on INT, as measured by rsfMRI, we identified the left AG as a key region whose atrophy induces its atypical intrinsic neural activity, which then appears to disturb neural dynamics within the overall DMN and consequently appears to contribute to cognitive impairment in AD, especially attention-related symptoms (Fig. 5). Given that INT length indicates the capacity of local information integration, these findings suggest that, in individuals with AD, the atrophy of the left AG affects the DMN’s capability to integrate a wide range of neural inputs to the network, which then appears to diminish efficient cognitive processing and leads to some of the AD symptoms, such as impaired attention control.

**Figure 5.**
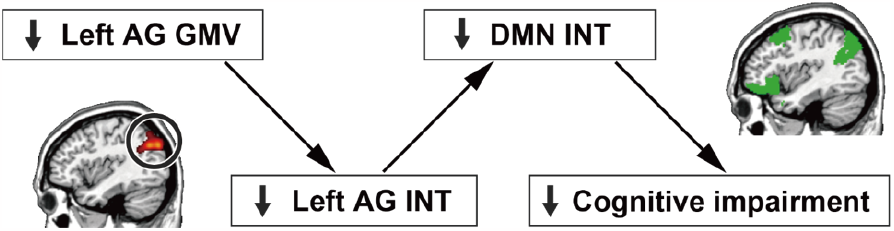
Links between neuroanatomical, functional and behavioural changes in AD. The two mediation analyses (Figs. 2G and 3B) suggest that the atypically smaller GMV of the left AG shortened its INT, and such shorter AG INT reduced INT of the DMN, which led to the cognitive impairment of AD.

The ostensible linkage between shorter DMN INT and AD symptoms is consistent with previous studies on the DMN in healthy humans. That is, several human neuroimaging studies reported that regions within the DMN are more likely to exhibit longer INT than those in sensory-related brain networks^29,55-57^, which in turn permits the DMN to pool and integrate neural inputs from other regions for complex behavioural and cognitive processes^38,41,55,56,58^. In particular, the DMN has a central role in orchestrating diverse cognitive functions in older individuals^25^. Given these previous reports, the current findings on the adverse effects of the atypically shorter INT of the DMN on cognitive capacities seem neurobiologically reasonable.

The crucial role of the AG in AD has been supported by outcomes of previous studies. In rsfMRI studies^59,60^, the AG showed atypically weak functional connectivity with other brain regions.

Research employing positron emission tomography reported hypometabolism, atrophy, and amyloid and tau deposition in parietal cortical regions, including the AG, in AD^26,61-64^. The current observation about the importance of the left AG in AD cognitive decline represents a key observation that fills a gap between AG anatomy and function and provides a more integrated view on the biological mechanisms underpinning AD.

Our findings suggest the possibility that the left AG could represent a target area for non-invasive brain stimulation to slow or even modify AD symptoms. Indeed, accumulating evidence shows the beneficial effects of repetitive transcranial magnetic stimulation (rTMS) on AD^65^: for example, a recent rTMS study demonstrated that stimulation of the precuneus could mitigate the cognitive decline seen in AD patients^66^. Considering our findings, a brain stimulation method to modify INT of the AG would be a new approach to alleviate the progressive cognitive impairments of the neurodegenerative dementia.

In addition, INT of the left AG could serve as a biomarker for AD, which also is relatively facile to measure. If the current finding were verified in a larger sample, a mere 15-min structural and functional MRI scan would enable evaluation of the degree of the neurodegeneration and resultant dysfunction of the DMN in individuals with AD^67^.

One limitation of the current study is that we used a cross-sectional dataset with a relatively limited AD sample size. Nevertheless, the mediation analyses that we conducted indicated direct links between local atrophy, local brain dynamics, network neural dynamics and behaviour (Fig. 5). However, longitudinal studies with larger-sized cohorts will be necessary for the direct examination of the causal effects of such alteration of the local brain activity on AD symptoms.

Another limitation concerns the absence of directly investigating pathological links between INT and amyloid and tau depositions, which was far beyond the scope of the employed data set. However, we confirmed the effects of the GMV reduction on the shorter INT in the left AG, and such a decrease in GMV is thought to reflect neuronal atrophy^68^. Future studies would be necessary to directly examine the relationships between INT and amyloid/tau depositions in individuals with AD.

## Data Availability

All data produced in the present study are available upon reasonable request to the authors.

https://www.oasis-brains.org/

## Acknowledgements

This work was supported by Grant-in-aid for Research Activity from Japan Society for Promotion of Sciences (SAM, 22K20879; TM, 22K07512; TW, 19H03535, 21H05679, 23H04217), AMED (TM, 22dk0207061s0101) and JST Moonshot R & D Program (TW, JPMJMS2021).

## References

1. Ballatore C, Lee VM, Trojanowski JQ. Tau-mediated neurodegeneration in Alzheimer’s disease and related disorders. Nat Rev Neurosci. Sep 2007;8(9):663–72. doi:10.1038/nrn2194

2. Braak H, Del Tredici K. The preclinical phase of the pathological process underlying sporadic Alzheimer’s disease. Brain. Oct 2015;138(Pt 10):2814–33. doi:10.1093/brain/awv236

3. Jack CR, Jr., Bennett DA, Blennow K, et al. NIA-AA Research Framework: Toward a biological definition of Alzheimer’s disease. Alzheimers Dement. Apr 2018;14(4):535–562. doi:10.1016/j.jalz.2018.02.018

4. Buckner RL, Snyder AZ, Shannon BJ, et al. Molecular, structural, and functional characterization of Alzheimer’s disease: evidence for a relationship between default activity, amyloid, and memory. J Neurosci. Aug 24 2005;25(34):7709–17. doi:10.1523/JNEUROSCI.2177-05.2005

5. Scholl M, Lockhart SN, Schonhaut DR, et al. PET Imaging of Tau Deposition in the Aging Human Brain. Neuron. Mar 2 2016;89(5):971–982. doi:10.1016/j.neuron.2016.01.028

6. Zott B, Busche MA, Sperling RA, Konnerth A. What Happens with the Circuit in Alzheimer’s Disease in Mice and Humans? Annu Rev Neurosci. Jul 8 2018;41:277–297. doi:10.1146/annurev-neuro-080317-061725

7. Sperling RA, Aisen PS, Beckett LA, et al. Toward defining the preclinical stages of Alzheimer’s disease: recommendations from the National Institute on Aging-Alzheimer’s Association workgroups on diagnostic guidelines for Alzheimer’s disease. Alzheimers Dement. May 2011;7(3):280–92. doi:10.1016/j.jalz.2011.03.003

8. Buckner RL, Andrews-Hanna JR, Schacter DL. The brain’s default network: anatomy, function, and relevance to disease. Ann N Y Acad Sci. Mar 2008;1124:1–38. doi:10.1196/annals.1440.011

9. Jagust W. Imaging the evolution and pathophysiology of Alzheimer disease. Nat Rev Neurosci. Nov 2018;19(11):687–700. doi:10.1038/s41583-018-0067-3

10. Thompson PM, Hayashi KM, de Zubicaray G, et al. Dynamics of gray matter loss in Alzheimer’s disease. J Neurosci. Feb 1 2003;23(3):994–1005. doi:10.1523/JNEUROSCI.23-03-00994.2003

11. Scahill RI, Schott JM, Stevens JM, Rossor MN, Fox NC. Mapping the evolution of regional atrophy in Alzheimer’s disease: unbiased analysis of fluid-registered serial MRI. Proc Natl Acad Sci U S A. Apr 2 2002;99(7):4703–7. doi:10.1073/pnas.052587399

12. Ossenkoppele R, Cohn-Sheehy BI, La Joie R, et al. Atrophy patterns in early clinical stages across distinct phenotypes of Alzheimer’s disease. Hum Brain Mapp. Nov 2015;36(11):4421–37. doi:10.1002/hbm.22927

13. Whitwell JL, Jack CR, Jr., Przybelski SA, et al. Temporoparietal atrophy: a marker of AD pathology independent of clinical diagnosis. Neurobiol Aging. Sep 2011;32(9):1531–41. doi:10.1016/j.neurobiolaging.2009.10.012

14. Seeley WW, Crawford RK, Zhou J, Miller BL, Greicius MD. Neurodegenerative diseases target large-scale human brain networks. Neuron. Apr 16 2009;62(1):42–52. doi:10.1016/j.neuron.2009.03.024

15. Tetreault AM, Phan T, Orlando D, et al. Network localization of clinical, cognitive, and neuropsychiatric symptoms in Alzheimer’s disease. Brain. Apr 1 2020;143(4):1249–1260. doi:10.1093/brain/awaa058

16. Greicius MD, Srivastava G, Reiss AL, Menon V. Default-mode network activity distinguishes Alzheimer’s disease from healthy aging: evidence from functional MRI. Proc Natl Acad Sci U S A. Mar 30 2004;101(13):4637–42. doi:10.1073/pnas.0308627101

17. Badhwar A, Tam A, Dansereau C, Orban P, Hoffstaedter F, Bellec P. Resting-state network dysfunction in Alzheimer’s disease: A systematic review and meta-analysis. Alzheimers Dement (Amst). 2017;8:73–85. doi:10.1016/j.dadm.2017.03.007

18. Zhou J, Greicius MD, Gennatas ED, et al. Divergent network connectivity changes in behavioural variant frontotemporal dementia and Alzheimer’s disease. Brain. May 2010;133(Pt 5):1352–67. doi:10.1093/brain/awq075

19. Jones DT, Knopman DS, Gunter JL, et al. Cascading network failure across the Alzheimer’s disease spectrum. Brain. Feb 2016;139(Pt 2):547–62. doi:10.1093/brain/awv338

20. He Y, Wang L, Zang Y, et al. Regional coherence changes in the early stages of Alzheimer’s disease: a combined structural and resting-state functional MRI study. Neuroimage. Apr 1 2007;35(2):488–500. doi:10.1016/j.neuroimage.2006.11.042

21. Grothe MJ, Teipel SJ, Alzheimer’s Disease Neuroimaging I. Spatial patterns of atrophy, hypometabolism, and amyloid deposition in Alzheimer’s disease correspond to dissociable functional brain networks. Hum Brain Mapp. Jan 2016;37(1):35–53. doi:10.1002/hbm.23018

22. Franzmeier N, Neitzel J, Rubinski A, et al. Functional brain architecture is associated with the rate of tau accumulation in Alzheimer’s disease. Nat Commun. Jan 17 2020;11(1):347. doi:10.1038/s41467-019-14159-1

23. Hoenig MC, Bischof GN, Seemiller J, et al. Networks of tau distribution in Alzheimer’s disease. Brain. Feb 1 2018;141(2):568–581. doi:10.1093/brain/awx353

24. Jones DT, Graff-Radford J, Lowe VJ, et al. Tau, amyloid, and cascading network failure across the Alzheimer’s disease spectrum. Cortex. Dec 2017;97:143–159. doi:10.1016/j.cortex.2017.09.018

25. Ezaki T, Sakaki M, Watanabe T, Masuda N. Age-related changes in the ease of dynamical transitions in human brain activity. Hum Brain Mapp. Jun 2018;39(6):2673–2688. doi:10.1002/hbm.24033

26. Putcha D, Eckbo R, Katsumi Y, Dickerson BC, Touroutoglou A, Collins JA. Tau and the fractionated default mode network in atypical Alzheimer’s disease. Brain Commun. 2022;4(2):fcac055. doi:10.1093/braincomms/fcac055

27. Watanabe T, Rees G, Masuda N. Atypical intrinsic neural timescale in autism. Elife. Feb 5 2019;8doi:10.7554/eLife.42256

28. Wengler K, Goldberg AT, Chahine G, Horga G. Distinct hierarchical alterations of intrinsic neural timescales account for different manifestations of psychosis. Elife. Oct 27 2020;9doi:10.7554/eLife.56151

29. Golesorkhi M, Gomez-Pilar J, Tumati S, Fraser M, Northoff G. Temporal hierarchy of intrinsic neural timescales converges with spatial core-periphery organization. Commun Biol. Mar 4 2021;4(1):277. doi:10.1038/s42003-021-01785-z

30. Hasson U, Yang E, Vallines I, Heeger DJ, Rubin N. A hierarchy of temporal receptive windows in human cortex. J Neurosci. Mar 5 2008;28(10):2539–50. doi:10.1523/JNEUROSCI.5487-07.2008

31. Lerner Y, Honey CJ, Silbert LJ, Hasson U. Topographic mapping of a hierarchy of temporal receptive windows using a narrated story. J Neurosci. Feb 23 2011;31(8):2906–15. doi:10.1523/JNEUROSCI.3684-10.2011

32. Stephens GJ, Honey CJ, Hasson U. A place for time: the spatiotemporal structure of neural dynamics during natural audition. J Neurophysiol. Nov 2013;110(9):2019–26. doi:10.1152/jn.00268.2013

33. Yeshurun Y, Nguyen M, Hasson U. Amplification of local changes along the timescale processing hierarchy. Proc Natl Acad Sci U S A. Aug 29 2017;114(35):9475–9480. doi:10.1073/pnas.1701652114

34. Chaudhuri R, Knoblauch K, Gariel MA, Kennedy H, Wang XJ. A Large-Scale Circuit Mechanism for Hierarchical Dynamical Processing in the Primate Cortex. Neuron. Oct 21 2015;88(2):419–31. doi:10.1016/j.neuron.2015.09.008

35. Honey CJ, Thesen T, Donner TH, et al. Slow cortical dynamics and the accumulation of information over long timescales. Neuron. Oct 18 2012;76(2):423–34. doi:10.1016/j.neuron.2012.08.011

36. Cavanagh SE, Wallis JD, Kennerley SW, Hunt LT. Autocorrelation structure at rest predicts value correlates of single neurons during reward-guided choice. Elife. Oct 5 2016;5doi:10.7554/eLife.18937

37. Gollo LL. Exploring atypical timescales in the brain. Elife. Feb 5 2019;8doi:10.7554/eLife.45089

38. Murray JD, Bernacchia A, Freedman DJ, et al. A hierarchy of intrinsic timescales across primate cortex. Nat Neurosci. Dec 2014;17(12):1661–3. doi:10.1038/nn.3862

39. Ogawa T, Komatsu H. Differential temporal storage capacity in the baseline activity of neurons in macaque frontal eye field and area V4. J Neurophysiol. May 2010;103(5):2433–45. doi:10.1152/jn.01066.2009

40. Runyan CA, Piasini E, Panzeri S, Harvey CD. Distinct timescales of population coding across cortex. Nature. Aug 3 2017;548(7665):92–96. doi:10.1038/nature23020

41. Hasson U, Chen J, Honey CJ. Hierarchical process memory: memory as an integral component of information processing. Trends Cogn Sci. Jun 2015;19(6):304–13. doi:10.1016/j.tics.2015.04.006

42. Gauthier B, Eger E, Hesselmann G, Giraud AL, Kleinschmidt A. Temporal tuning properties along the human ventral visual stream. J Neurosci. Oct 10 2012;32(41):14433–41. doi:10.1523/JNEUROSCI.2467-12.2012

43. Marcus DS, Wang TH, Parker J, Csernansky JG, Morris JC, Buckner RL. Open Access Series of Imaging Studies (OASIS): cross-sectional MRI data in young, middle aged, nondemented, and demented older adults. J Cogn Neurosci. Sep 2007;19(9):1498–507. doi:10.1162/jocn.2007.19.9.1498

44. LaMontagne PJ, Benzinger TLS, Morris JC, et al. OASIS-3: Longitudinal Neuroimaging, Clinical, and Cognitive Dataset for Normal Aging and Alzheimer Disease. MedRxiv. 2019;doi:10.1101/2019.12.13.19014902

45. Morris JC, Weintraub S, Chui HC, et al. The Uniform Data Set (UDS): clinical and cognitive variables and descriptive data from Alzheimer Disease Centers. Alzheimer Dis Assoc Disord. Oct-Dec 2006;20(4):210–6. doi:10.1097/01.wad.0000213865.09806.92

46. Morris JC, Storandt M, Miller JP, et al. Mild cognitive impairment represents early-stage Alzheimer disease. Arch Neurol. Mar 2001;58(3):397–405. doi:10.1001/archneur.58.3.397

47. Morris JC. The Clinical Dementia Rating (CDR): current version and scoring rules. Neurology. 1993;

48. Marcus DS, Fotenos AF, Csernansky JG, Morris JC, Buckner RL. Open access series of imaging studies: longitudinal MRI data in nondemented and demented older adults. J Cogn Neurosci. Dec 2010;22(12):2677–84. doi:10.1162/jocn.2009.21407

49. Hanseeuw BJ, Betensky RA, Mormino EC, et al. PET staging of amyloidosis using striatum. Alzheimers Dement. Oct 2018;14(10):1281–1292. doi:10.1016/j.jalz.2018.04.011

50. Konijnenberg E, Tijms BM, Gobom J, et al. APOE epsilon4 genotype-dependent cerebrospinal fluid proteomic signatures in Alzheimer’s disease. Alzheimers Res Ther. May 27 2020;12(1):65. doi:10.1186/s13195-020-00628-z

51. Yeo BT, Krienen FM, Sepulcre J, et al. The organization of the human cerebral cortex estimated by intrinsic functional connectivity. J Neurophysiol. Sep 2011;106(3):1125–65. doi:10.1152/jn.00338.2011

52. Ashburner J, Friston KJ. Unified segmentation. Neuroimage. Jul 1 2005;26(3):839–51. doi:10.1016/j.neuroimage.2005.02.018

53. Ashburner J. A fast diffeomorphic image registration algorithm. Neuroimage. Oct 15 2007;38(1):95–113. doi:10.1016/j.neuroimage.2007.07.007

54. Weintraub S, Salmon D, Mercaldo N, et al. The Alzheimer’s Disease Centers’ Uniform Data Set (UDS): the neuropsychologic test battery. Alzheimer Dis Assoc Disord. Apr-Jun 2009;23(2):91–101. doi:10.1097/WAD.0b013e318191c7dd

55. Wolff A, Berberian N, Golesorkhi M, Gomez-Pilar J, Zilio F, Northoff G. Intrinsic neural timescales: temporal integration and segregation. Trends Cogn Sci. Feb 2022;26(2):159–173. doi:10.1016/j.tics.2021.11.007

56. Golesorkhi M, Gomez-Pilar J, Zilio F, et al. The brain and its time: intrinsic neural timescales are key for input processing. Commun Biol. Aug 16 2021;4(1):970. doi:10.1038/s42003-021-02483-6

57. Ito T, Hearne LJ, Cole MW. A cortical hierarchy of localized and distributed processes revealed via dissociation of task activations, connectivity changes, and intrinsic timescales. Neuroimage. Nov 1 2020;221:117141. doi:10.1016/j.neuroimage.2020.117141

58. Cavanagh SE, Hunt LT, Kennerley SW. A Diversity of Intrinsic Timescales Underlie Neural Computations. Front Neural Circuits. 2020;14:615626. doi:10.3389/fncir.2020.615626

59. Liu Y, Yu C, Zhang X, et al. Impaired long distance functional connectivity and weighted network architecture in Alzheimer’s disease. Cereb Cortex. Jun 2014;24(6):1422–35. doi:10.1093/cercor/bhs410

60. Agosta F, Pievani M, Geroldi C, Copetti M, Frisoni GB, Filippi M. Resting state fMRI in Alzheimer’s disease: beyond the default mode network. Neurobiol Aging. Aug 2012;33(8):1564–78. doi:10.1016/j.neurobiolaging.2011.06.007

61. Lowe VJ, Wiste HJ, Senjem ML, et al. Widespread brain tau and its association with ageing, Braak stage and Alzheimer’s dementia. Brain. Jan 1 2018;141(1):271–287. doi:10.1093/brain/awx320

62. La Joie R, Perrotin A, Barre L, et al. Region-specific hierarchy between atrophy, hypometabolism, and beta-amyloid (Abeta) load in Alzheimer’s disease dementia. J Neurosci. Nov 14 2012;32(46):16265–73. doi:10.1523/JNEUROSCI.2170-12.2012

63. Chen Y, Wolk DA, Reddin JS, et al. Voxel-level comparison of arterial spin-labeled perfusion MRI and FDG-PET in Alzheimer disease. Neurology. Nov 29 2011;77(22):1977–85. doi:10.1212/WNL.0b013e31823a0ef7

64. Frings L, Hellwig S, Spehl TS, et al. Asymmetries of amyloid-beta burden and neuronal dysfunction are positively correlated in Alzheimer’s disease. Brain. Oct 2015;138(Pt 10):3089–99. doi:10.1093/brain/awv229

65. Chou YH, Ton That V, Sundman M. A systematic review and meta-analysis of rTMS effects on cognitive enhancement in mild cognitive impairment and Alzheimer’s disease. Neurobiol Aging. Feb 2020;86:1–10. doi:10.1016/j.neurobiolaging.2019.08.020

66. Koch G, Casula EP, Bonni S, et al. Precuneus magnetic stimulation for Alzheimer’s disease: a randomized, sham-controlled trial. Brain. Nov 21 2022;145(11):3776–3786. doi:10.1093/brain/awac285

67. Badhwar A, McFall GP, Sapkota S, et al. A multiomics approach to heterogeneity in Alzheimer’s disease: focused review and roadmap. Brain. May 1 2020;143(5):1315–1331. doi:10.1093/brain/awz384

68. Matsuda H. MRI morphometry in Alzheimer’s disease. Ageing Res Rev. Sep 2016;30:17–24. doi:10.1016/j.arr.2016.01.003

